# Does a waiting room increase same-day treatment for sexually transmitted infections among pregnant women? A quality improvement study at South African primary healthcare facilities

**DOI:** 10.1101/2025.02.12.25322145

**Authors:** Ranjana M.S. Gigi, Mandisa M. Mdingi, Lukas Bütikofer, Chibuzor M. Babalola, Jeffrey D. Klausner, Andrew Medina-Marino, Christina A. Muzny, Christopher M. Taylor, Janneke H.H.M. van de Wijgert, Remco P.H. Peters, Nicola Low

## Abstract

**Background:** Same-day testing and treatment of curable sexually transmitted infections (STI) is a strategy to reduce infection duration and onward transmission. South African primary healthcare facilities often lack sufficient waiting spaces. This study aimed to assess the proportion of, and factors influencing, pregnant women waiting for on-site STI test results before and after the installation of clinic-based waiting rooms.

**Methods:** We conducted an observational quality improvement study at 5 public primary healthcare facilities in South Africa from March 2021 to May 2023. The intervention was the installation of a waiting room in two clinics. Three clinics were used as comparators: two already had a waiting room in an existing building and one had access to a shared waiting area. The outcome was the percentage of women who waited for their STI test results. We conducted univariable and multivariable analyses and report marginal risk differences (with 95% confidence intervals, CI) of the proportions of women who waited for results. A subset of women answered structured questions about factors influencing their decision to wait for results.

**Results:** We analysed data from 624 women across the 5 facilities. Overall, 36% (95% CI 31, 40) waited for their test results (range 7% to 89%). In the two intervention clinics, 17% (95% CI 11, 24) waited for results before the introduction of a waiting room and 10% (95% CI 5, 18) after (crude absolute difference -7% (95% CI -16, +3), adjusted difference, -6% (95% CI -17, +5)). The percentages of pregnant women waiting for sexually transmitted infection test results were higher throughout the study period in 2 clinics which always had a dedicated waiting room than in 2 clinics where a waiting room was installed, or in 1 clinic, which only had access to a shared waiting area. Most women reported before testing that they did not intend to wait and none of the suggested factors would change their decision.

**Conclusions:** Introduction of a waiting room did not increase the proportion of women who waited for their results in this observational study. Future studies should investigate infrastructure, individual and test-based factors that affect same-day STI testing and treatment.

## Introduction

Antenatal clinic services should provide timely treatment for women with newly diagnosed conditions in pregnancy. In South Africa, approximately one-third of women attending antenatal clinics have been found to have at least one curable sexually transmitted infection, including *Chlamydia trachomatis*, *Neisseria gonorrhoeae* or *Trichomonas vaginalis* [1, 2]. In settings lacking laboratory STI diagnostic testing, women with vaginal discharge often receive syndromic management - an empiric approach in which people are treated for the most common potential causes [3]. Clinical management of STIs in pregnancy with rapid accurate diagnostic tests at the point of care would allow same-day targeted treatment [4], reduce overtreatment of symptomatic women without infection and detect asymptomatic infections [5, 6]. The GeneXpert platform (Cepheid, Sunnyvale, California) uses highly sensitive and specific nucleic amplification technology, which can detect *C. trachomatis, N. gonorrhoeae* and *T. vaginalis* within 60 to 90 minutes but requires an electricity supply.

The proportion of women who wait for results or receive same-day treatment for STIs detected on the GeneXpert platform in community or primary care settings in resource-limited settings, including sub-Saharan Africa has varied from 2% to 92% [7-12]. In a study in Zimbabwe, researchers investigated why only 2% (1/61) of adolescents received results on the same day after being tested in non-clinical community settings. Clinic staff reported delays resulting from workflow difficulties and power cuts, and that patients could not wait 90 minutes for results [7]. In our South African research project, called “Philani Ndiphile”, 39% of 125 pregnant women who tested positive for STIs using GeneXpert between March 2021 and March 2022, received same-day treatment. This percentage varied from 26% to 96% among the four clinics [13]. We observed that the clinic where most women waited for results had a comfortable designated waiting room.

The ‘Ideal Clinic’ initiative of the South African National Department of Health aims to create healthcare facilities with good infrastructure, including adequate waiting space and waiting times [14]. However, most healthcare facilities in South Africa need renovation and extension [14, 15]. In addition, the increasing frequency of planned power cuts (known as ‘load shedding’) could increase the turnaround time, from arrival at the clinic to receiving of test results [4, 16], in research settings where the GeneXpert test platform is available. In September 2022, the Philani Ndiphile project investigators planned to upgrade the physical conditions in two clinics by installing a waiting room. This decision provided an opportunity to observe whether there were differences in the percentages of pregnant women waiting for GeneXpert STI test results, according to the presence of a dedicated waiting room. The objectives of this study were to: 1) observe the percentage of women waiting for STI test results before and after installing a dedicated waiting room in two clinics; 2) compare percentages of women waiting for results in clinics with and without a waiting room; and 3) describe reasons why pregnant women did not intend to wait for results.

## Methods

We used the Standards for Quality Improvement Reporting Excellence (version 2.0) guidelines [17] to report this study. The statistical analysis plan is at: https://osf.io/zk4xd/.

### Study design and setting

This healthcare quality improvement study used an observational design. It was nested within the Philani Ndiphile research project, which aims to evaluate different screening strategies to decrease the burden of STIs among pregnant women and reduce adverse birth outcomes in Buffalo City Metropolitan Health District, Eastern Cape Province, South Africa [18, 19]. Enrolment into Philani Ndiphile commenced in the antenatal clinics of four public healthcare facilities between March and May 2021, with a fifth clinic starting in June 2022 [13]. We used study data collected from Philani Ndiphile participants from 29 March 2021 to 12 May 2023.

Clinics A, B and E are community healthcare centres, and Clinics C and D primary healthcare facilities, with the former being larger and offering more services than the latter (Table 1).^1^ Clinics A and E are in peri-urban townships. Clinic A is in a more densely populated area with mainly informal dwellings than Clinic E with a mix of formal and informal dwellings. Clinic B is in a larger peri-urban township with mainly formal dwellings. Clinic D is in a semi-rural area with formal dwellings. Clinic C is in a sparsely populated peri-urban town with formal dwellings.

**Table 1.** Description of the clinic settings and research sites at baseline.

|  | Intervention clinics |  | Non-intervention clinics |  |  |
| --- | --- | --- | --- | --- | --- |
| Clinic<sup>1</sup> | Clinic A | Clinic B | Clinic C | Clinic D | Clinic E |
| Description of area |  |  |  |  |  |
| Type of health facility | Community health centre | Community health centre | Primary healthcare | Primary healthcare | Community health centre |
| Type of settlement | peri-urban township, informal dwellings | peri-urban township, formal dwellings | peri-urban town, formal dwellings | semi-rural area, formal dwellings | peri-urban township, informal and formal dwellings |
| Population density (per km<sup>2</sup>) [20] | 17410 | 3443 | 519 | 1796 | 4226 |
| Distance to the nearest shop selling food (metres) | 500 | 600 | 300 | 80 | 550 |
| Description of research site |  |  |  |  |  |
| Site description | 14m<sup>2</sup> metal freight container accessible through the backdoor of the healthcare facility | 14m<sup>2</sup> metal freight container in the parking lot of the healthcare facility, across from the main entrance | 3-room area inside the healthcare facility building | 4-room free-standing building within the grounds of the healthcare facility | One small room inside the healthcare facility, shared with other antenatal services |
| Water dispenser for participants | Yes | Yes | Yes | Yes | No |
| Bathroom | Inside the facility | Inside the facility | Across the courtyard | Across from the waiting room | In the same passage |
| Waiting room description | No dedicated waiting room at project start, camp chairs in an uncovered area, no privacy, no protection from the weather | No dedicated waiting room at project start, camp chairs in an uncovered area, limited privacy, very busy area, no protection from weather | Dedicated waiting room with chairs and a couch, private, not visible from the rest of the facility, cool in summer but cold in winter | Dedicated waiting room with chairs and a couch, private, not visible from the rest of the facility, hot in summer and cold in winter | No dedicated waiting room throughout, bench in the general waiting area, limited privacy, busy area, cool in summer, cold in winter |
| Research nurse clinical experience, years | 9 | 9 | 15 | 33 | 9 |

1. Clinic identification labels A to E do not correspond with labels reported in Mdingi MM, et al. [13]

The installation of a dedicated waiting room (described as the intervention) was implemented at Clinics A and B (Table 1, Figure 1). At these clinics, the Philani Ndiphile research procedures were initially conducted out of 14m^2^ (approximately 5.9m x 2.3m) metal freight containers on the community healthcare centre grounds and participants had to wait outside in an uncovered area. At Clinics C and D, research procedures were conducted in multiple rooms inside the clinic building. Only study participants could use these dedicated comfortable waiting rooms. At Clinic E, research study staff shared a small room with other antenatal care services inside the clinic building. Participants used the same waiting area, which was a corridor, as the regular antenatal clinic attendees. We judged this facility to be sub-optimal for an ‘Ideal Clinic’ [14]. Since no upgrade was planned, we considered it separately as having ‘no dedicated waiting area’.

**Figure 1.**
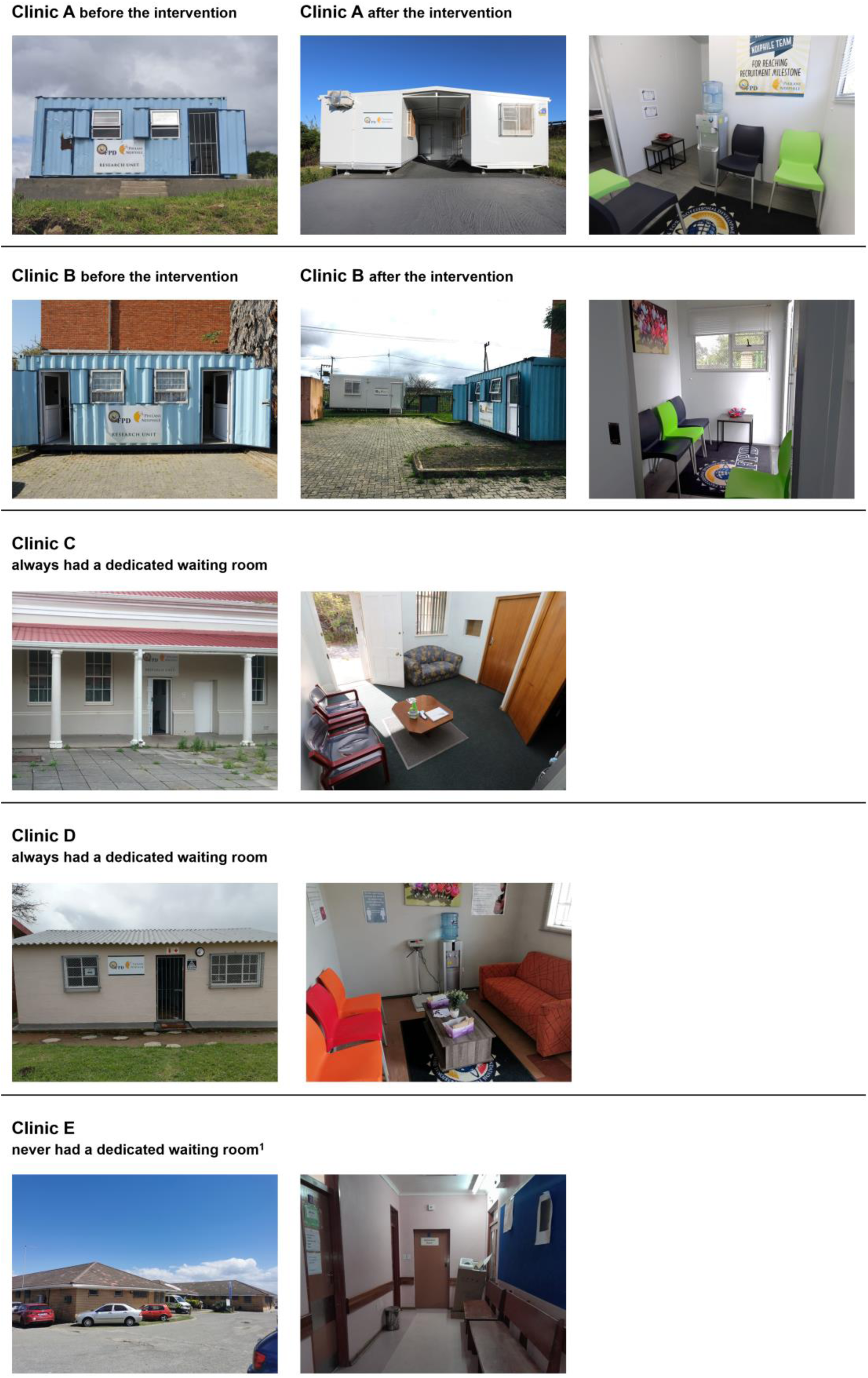
Clinics before and after the intervention. Legend: 1, waiting area shared with regular clinic patients

### Study population

At their enrolment visit, Philani Ndiphile study participants who received same-day STI testing according to the study protocol in the clinics, were included in this study. Participants were aged ≥18 years and <27 weeks of gestation at enrolment. A research nurse conducted a physical examination and collected vaginal swabs as early as possible during the enrolment visit in all clinics. One swab was tested on-site on the GeneXpert machine. The processing time for the combined assay for *C. trachomatis* and *N. gonorrhoeae* is 90 minutes and for the assay for *T. vaginalis* 60 minutes; the tests run in parallel on the same machine. Times from arrival at the clinic until testing and from the end of processing until receipt of results were not measured.

### Intervention

Clinics A and B were provided with a new air-conditioned 18m^2^ (approximately 5.9m x 3.0m) container, with one room furnished as a waiting room with seating space, a water dispenser, and a separate bathroom (Table 1, Figure 1). At Clinic A, the original 14m^2^ freight container was also replaced. A newly installed roof between the two new containers could also be used as an additional outside covered waiting area. At Clinic B, the 14m^2^ freight container remained. The intervention, introduction of the new waiting rooms, started operating on 14^th^ December 2022 (Figure 2).

**Figure 2.**
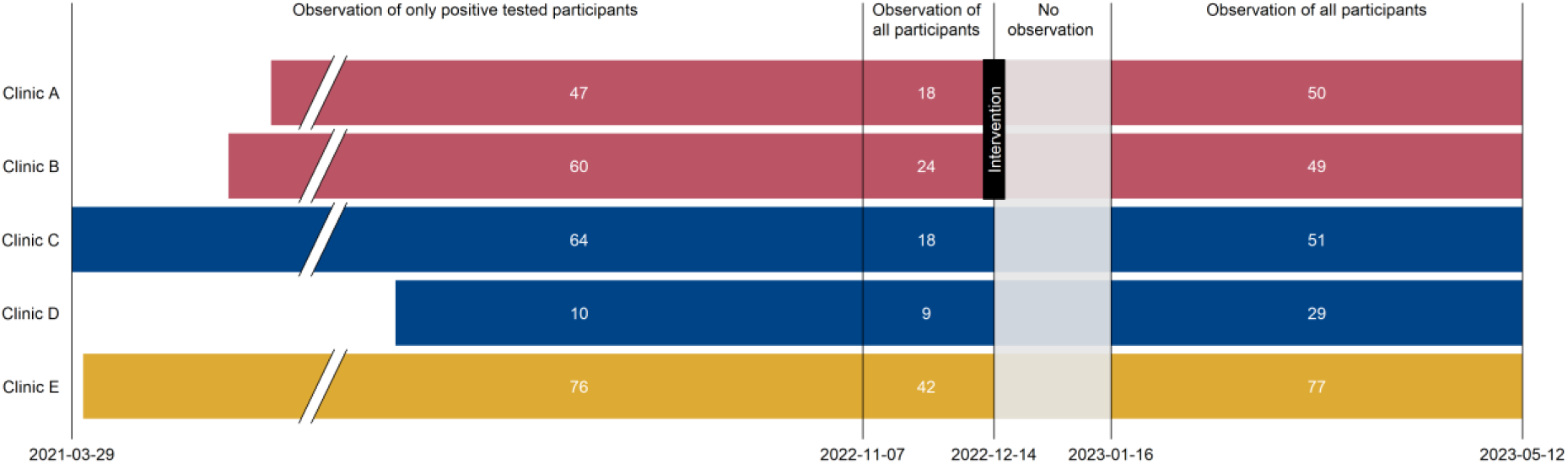
Study procedures from 29 March 2021 to 12 May 2023, by clinic. Legend: Clinics A and B (red bars; intervention clinics) initially had no dedicated waiting room; new waiting rooms were implanted in December 2022. Clinics C and D (blue bars; control clinics group 1) always had dedicated waiting rooms. Clinic E (yellow bar, control clinic group 2) never had a dedicated waiting room and did not receive any intervention; control group 2. Break in x-axis from 01 June 2021 to 01 June 2022. White numbers in each bar are numbers of participants for each time period and clinic.

### Outcomes

The primary outcome was the percentage of participants who waited to receive their STI test results on the same day. The secondary outcome was the response to structured questions about reasons for waiting for same-day STI test results.

### Sample size

We used study data about all STI tests and the proportions of participants waiting for results in the Clinics A and B before the intervention (26%, 17/65 and 10%, 8/84). Guided by the proportion of participants waiting in Clinic C (94%, 77/82) which always had a waiting room, we considered a conservative increase to 45%. Based on 26% of participants waiting and 147 participants with complete pre-intervention data, a total of 220 participants would result in more than 0.8 power to detect an improvement to 45% at an alpha of 0.05, based on a chi-squared test. We aimed to observe at least 80 participants post-intervention in Clinics A and B combined.

### Data collection

We recorded data in a Research Electronic Data Capture software (REDCap, Vanderbilt University, Tennessee, USA) project database [21]. From 29 March 2021 to 6 November 2022, we only collected the dates of testing and treatment for participants with a positive STI result (Figure 2). We classified the participant as having waited for her result if the dates of testing and treatment were identical. From 7 November 2022, we captured data from all participants, irrespective of their STI test result. The first month after the waiting rooms were installed on 14 December 2022 coincided with a holiday period. We considered this a ‘settling in’ period for the intervention and did not analyse data for this period (Figure 2). From 14 September 2022 onwards, all participants from all clinics were asked structured questions about their intention to wait, what would make them wait and, for participants who did not intend to wait but actually received same-day results, what were their reasons. We obtained national-level data about load shedding from the national power supplier. We applied the national level of load shedding that was in operation at midday of each day in the observation period to each clinic. We defined three groups: no load shedding, stages 1-3 (3-6 hours of power outage per day), and stages 4-6 (6-12 hours of power outage per day) and added this information to the database at the participant level.

### Statistical analysis

We summarised continuous baseline characteristics using means and standard deviations, or medians and interquartile ranges, and categorical variables as frequencies. We compared baseline characteristics of participants included in the analyses with all those eligible for same-day STI testing, using Mann-Whitney or chi-square tests.

Objective 1: we calculated the percentages (with 95% confidence intervals, CI) of women who waited for results (number who waited for their results divided by number tested), by clinic and intervention group. We made run charts, which display monthly aggregated percentages, by clinic and intervention status [22]. Objective 2: To analyse the primary outcome for each woman (waited for results, yes/no), we used binomial generalised estimating equations, with an independence correlation structure and the clinic as a cluster variable. We pre-specified 6 explanatory variables, which could have influenced waiting. Participant factors were employment status, symptom status and STI test positivity. Although the result is not known at the time of testing, women who are aware of sexual practices that increase STI acquisition risk or women who have symptoms might be more likely to wait for results under any circumstances [7, 23]. Clinic setting factors were the distance to the nearest food shop, which was a proxy for clinic location and ability to buy food whilst waiting, years of clinical experience of the research nurse and level of electrical load shedding. The primary analysis model compared data for women attending post-intervention (waiting room available) vs. pre-intervention (no waiting room) for Clinics A and B only. We conducted two secondary analyses, using data for all clinics. In the first model, waiting room was a binary variable with each woman categorised as attending a clinic in which a waiting room was available vs. not available. In the second model, waiting room was a categorical variable: never available, available from the beginning, and waiting room introduced. We conducted multivariable analyses and planned to adjust for all pre-specified explanatory variables in each model. We also conducted a subgroup analysis using data from Clinics A and B, according to the STI test, to investigate whether women who might have thought they had an STI were more likely to wait for their results under any circumstances. The model included an interaction term for STI and the intervention as additional covariates. We report results from each statistical model as the marginal risk difference (with 95% CI). Objective 3: we tabulated responses to structured questions about factors that might make the participant wait or change her mind and report these as frequencies and percentages.

### Ethical considerations

All participants provide written informed consent to participate in the Philani Ndiphile study. The Philani Ndiphile study received approval from the University of Cape Town’s Human Research Ethics Committee (Reference: 676/2019) and from the local Department of Health (Reference: EC_202010_017). The study reported here is a healthcare quality improvement study, which did not require ethical committee review. Authorisation to analyse de-identified data at the University of Bern has been granted by the Canton of Bern Ethics Committee (Reference 2021-01209).

## Results

From 29 March 2021 to 12 May 2023, a total of 1986 participants were enrolled into the Philani Ndiphile study and 1500 participants received same-day testing, according to study protocol (Figure 3). We excluded data from 822 participants enrolled before 7 November 2022, because we did not initially collect data about waiting for those with a negative STI test result, and from 54 participants during the settling in (no observation) period. Our final dataset included 624 participants: 257 in the period during which we only collected waiting data on participants with a positive STI test and 367 in the period during which we collected waiting data on all participants (Figure 2). Of these, 99 participants were enrolled post-intervention in Clinics A and B combined. There were no substantial differences in socio-economic and health characteristics of included and excluded participants (Additional File 1).

**Figure 3.**
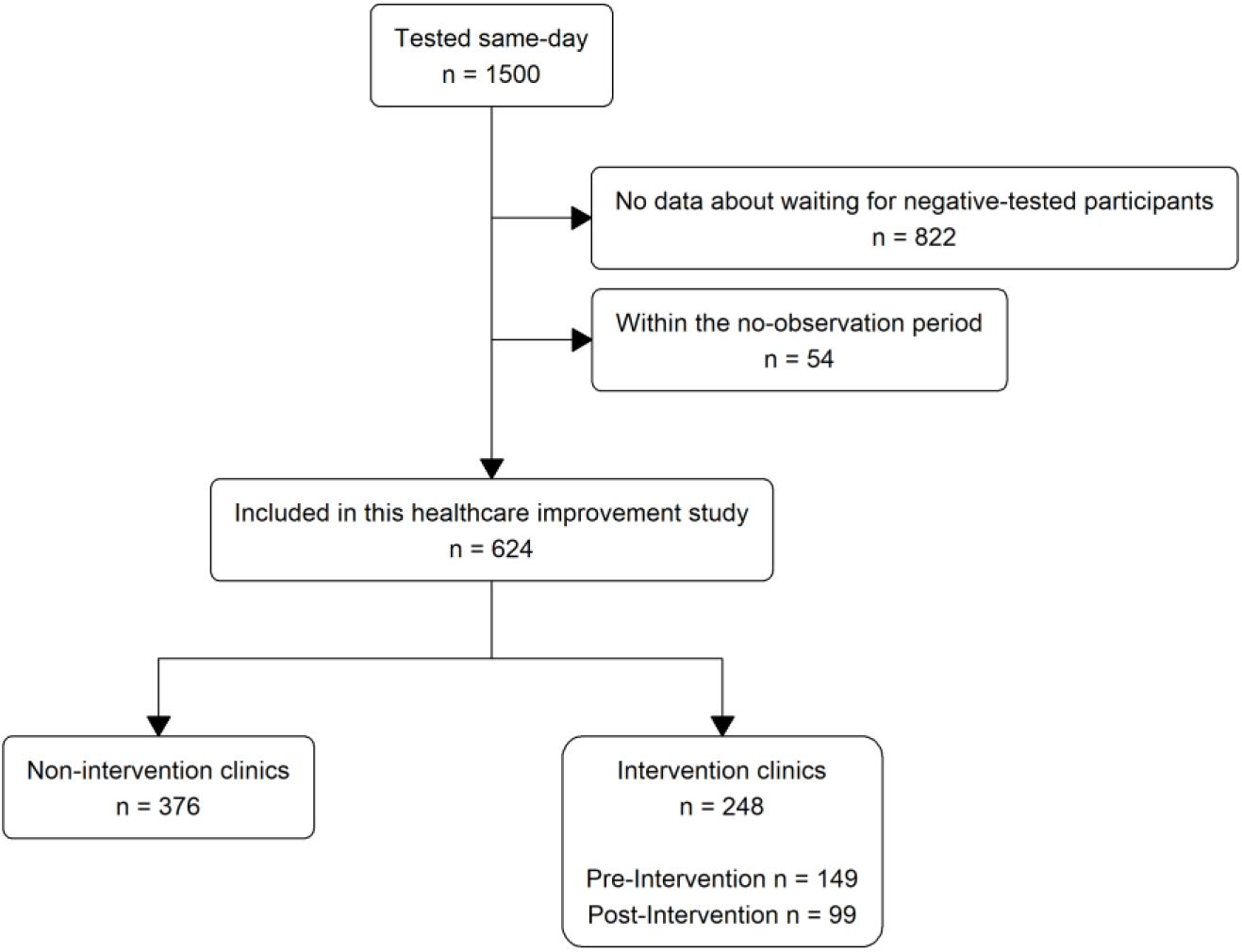
Flow chart of study participant selection.

The characteristics of included participants in each clinic are shown in Table 2. Across the 5 clinics, the median age was 27 years, and 31% of women were living with HIV. Participants reported waiting from around half an hour to two hours to see a public health nurse or a doctor. Research nurses in Clinics A, B and E had the same level of clinical experience. For these clinics, the population density and distance to the nearest shop were also very similar (Table 1).

**Table 2.**
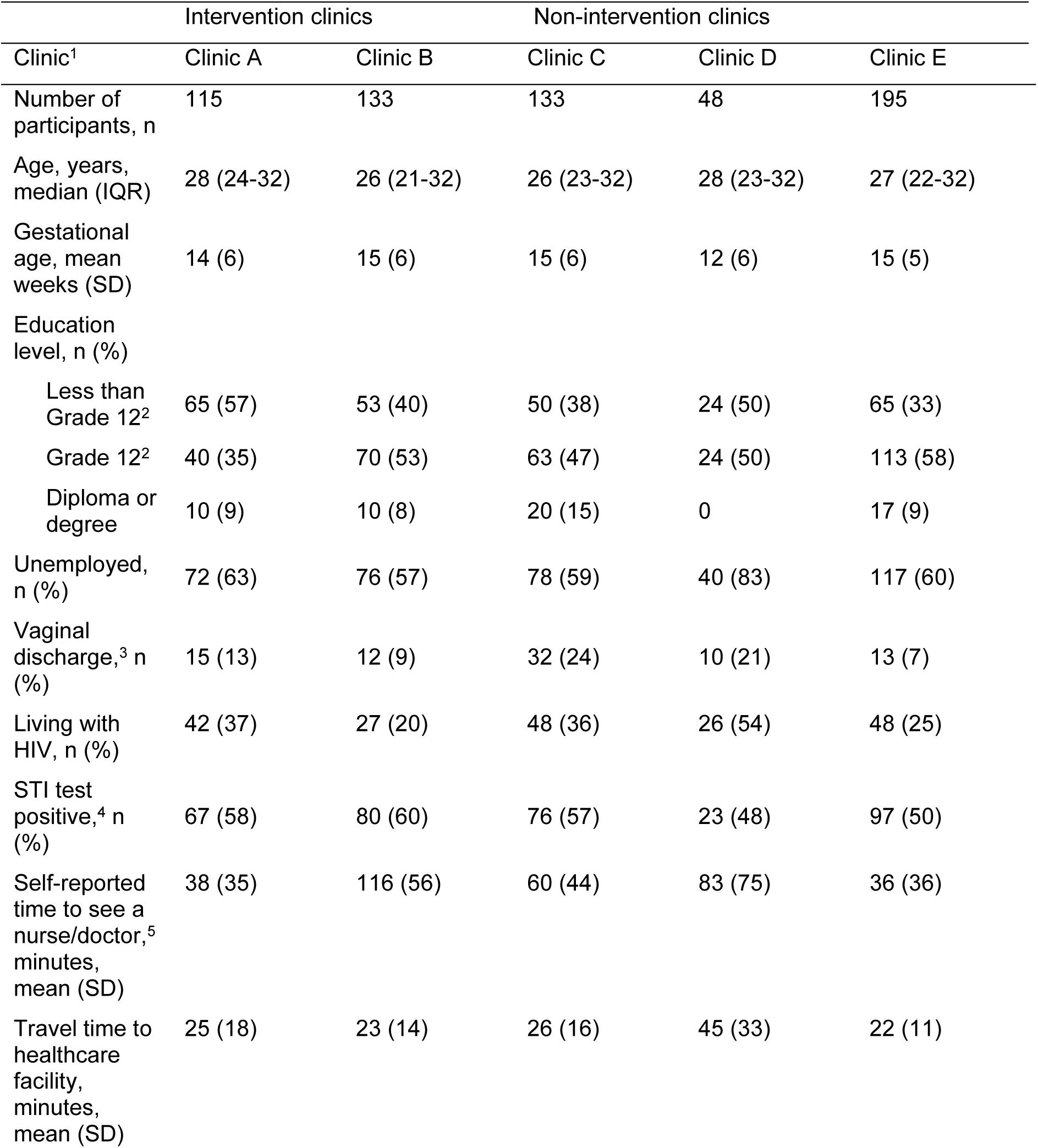
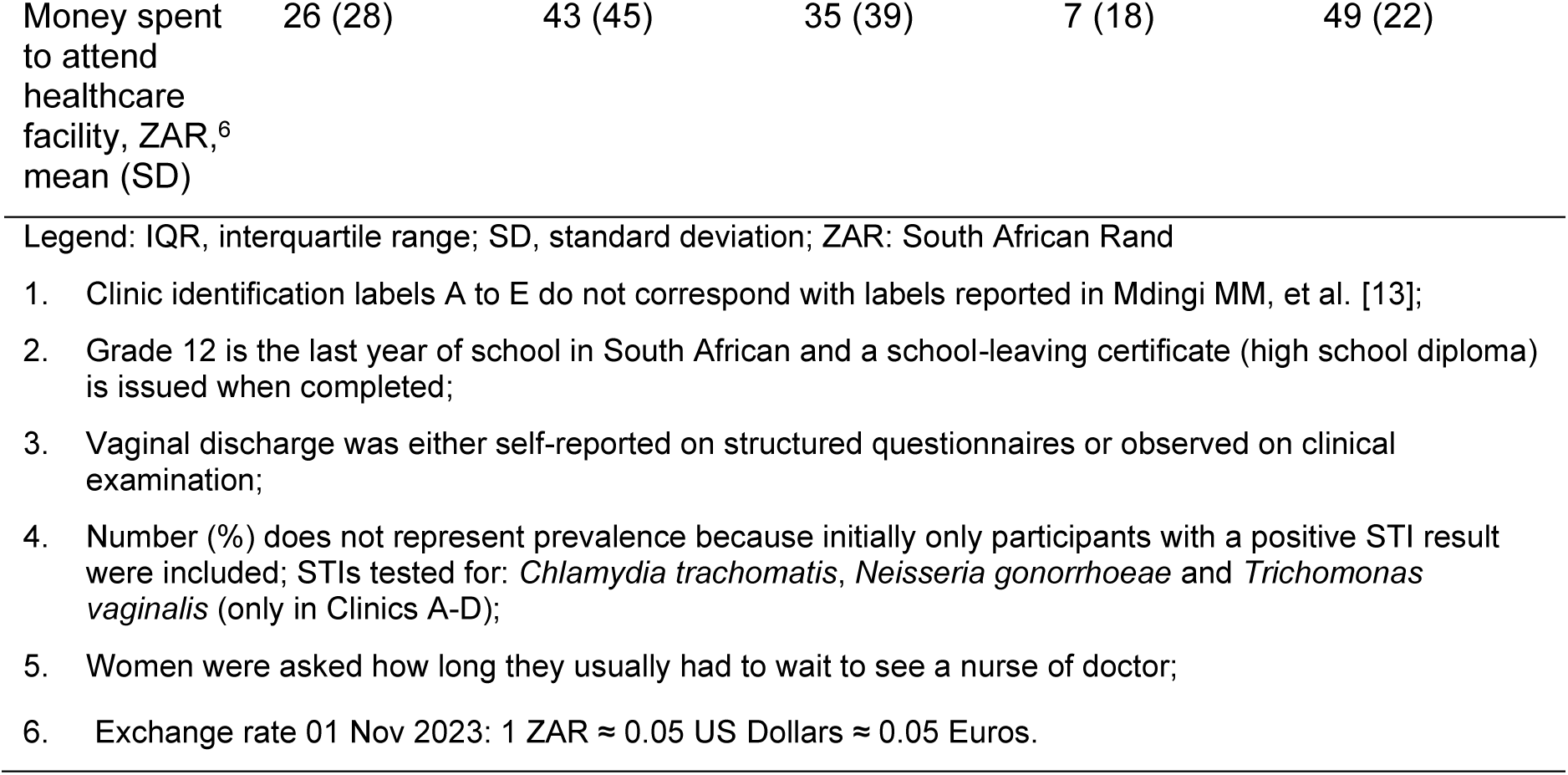
Baseline characteristics of included participants by clinic.

|  | Intervention clinics |  | Non-intervention clinics |  |  |
| --- | --- | --- | --- | --- | --- |
| Clinic<sup>1</sup> | Clinic A | Clinic B | Clinic C | Clinic D | Clinic E |
| Number of participants, n | 115 | 133 | 133 | 48 | 195 |
| Age, years, median (IQR) | 28 (24-32) | 26 (21-32) | 26 (23-32) | 28 (23-32) | 27 (22-32) |
| Gestational age, mean weeks (SD) | 14 (6) | 15 (6) | 15 (6) | 12 (6) | 15 (5) |
| Education level, n (%) |  |  |  |  |  |
| Less than Grade 12<sup>2</sup> | 65 (57) | 53 (40) | 50 (38) | 24 (50) | 65 (33) |
| Grade 12<sup>2</sup> | 40 (35) | 70 (53) | 63 (47) | 24 (50) | 113 (58) |
| Diploma or degree | 10 (9) | 10 (8) | 20 (15) | 0 | 17 (9) |
| Unemployed, n (%) | 72 (63) | 76 (57) | 78 (59) | 40 (83) | 117 (60) |
| Vaginal discharge,<sup>3</sup> n (%) | 15 (13) | 12 (9) | 32 (24) | 10 (21) | 13 (7) |
| Living with HIV, n (%) | 42 (37) | 27 (20) | 48 (36) | 26 (54) | 48 (25) |
| STI test positive,<sup>4</sup> n (%) | 67 (58) | 80 (60) | 76 (57) | 23 (48) | 97 (50) |
| Self-reported time to see a nurse/doctor,<sup>5</sup> minutes, mean (SD) | 38 (35) | 116 (56) | 60 (44) | 83 (75) | 36 (36) |
| Travel time to healthcare facility, minutes, mean (SD) | 25 (18) | 23 (14) | 26 (16) | 45 (33) | 22 (11) |
| Money spent to attend healthcare facility, ZAR,<sup>6</sup> mean (SD) | 26 (28) | 43 (45) | 35 (39) | 7 (18) | 49 (22) |

Legend: IQR, interquartile range; SD, standard deviation; ZAR: South African Rand
1. Clinic identification labels A to E do not correspond with labels reported in Mdingi MM, et al. [13]; 2. Grade 12 is the last year of school in South African and a school-leaving certificate (high school diploma) is issued when completed; 3. Vaginal discharge was either self-reported on structured questionnaires or observed on clinical examination; 4. Number (%) does not represent prevalence because initially only participants with a positive STI result were included; STIs tested for: *Chlamydia trachomatis*, *Neisseria gonorrhoeae* and *Trichomonas vaginalis* (only in Clinics A-D); 5. Women were asked how long they usually had to wait to see a nurse or doctor; 6. Exchange rate 01 Nov 2023: 1 ZAR ≈ 0.05 US Dollars ≈ 0.05 Euros.

The monthly run chart (Figure 4) shows that the percentages of women waiting for their test results before and after the intervention did not change substantially and that the percentage waiting in Clinics C and D, which always had dedicated waiting room, remained higher throughout the study period than in the intervention Clinics A and B and in Clinic E, which never had a dedicated waiting room. Summary data for each clinic separately are shown in Additional File 2.

**Figure 4.**
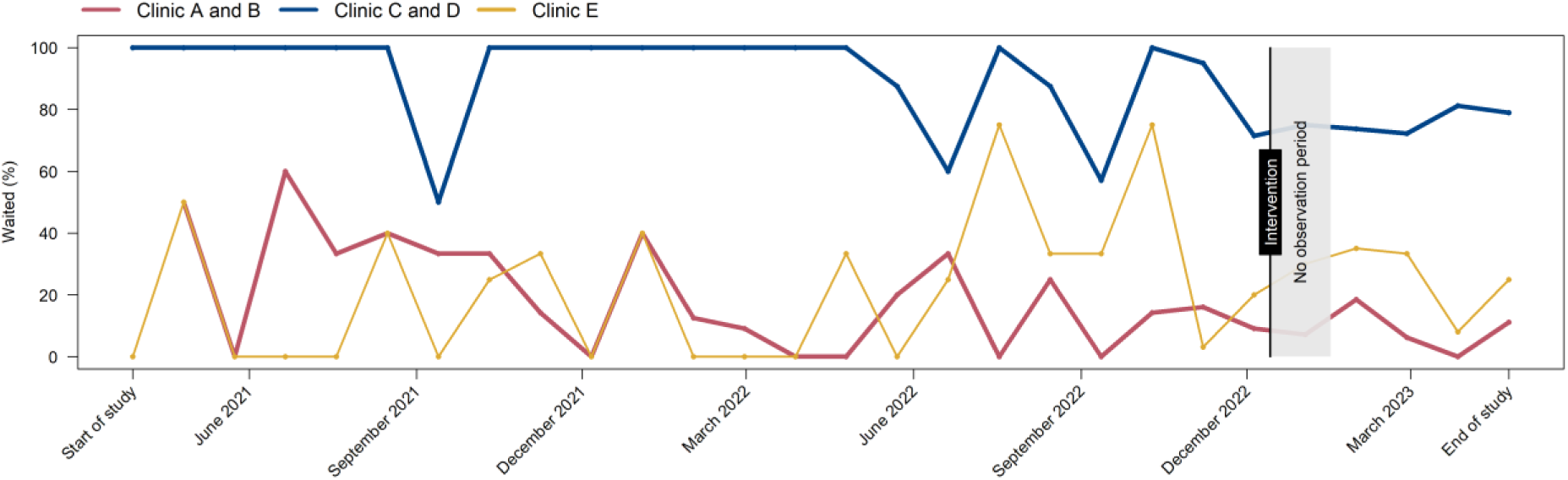
Monthly run chart showing the percentage of participants waiting over time, by clinic and intervention. Legend: Clinics A and B (red) intervention clinics; Clinics C and D (blue) always had a dedicated waiting room, control group 1; Clinic E (yellow) never had a dedicated waiting, control group 2. Intervention start was 14 December 2022; no observation from 14 December 2022 to 16 January 2023.

In the primary analysis, for the intervention clinics, Clinics A and B combined, 17% (CI 11 to 24) of participants waited for their result before the intervention and 10% (CI 5 to 18) after the addition of a waiting room (absolute difference, -7% (CI -16 to +3) (Table 3). After adjustment for employment status, STI test positivity, symptoms, load shedding and distance to the nearest shop, the absolute adjusted percentage waiting was 6% (CI -16 to +2) lower after the intervention (Table 3). The years of research nurse experience could not be included in the model because the values of the variable were identical in both clinics.

**Table 3.**
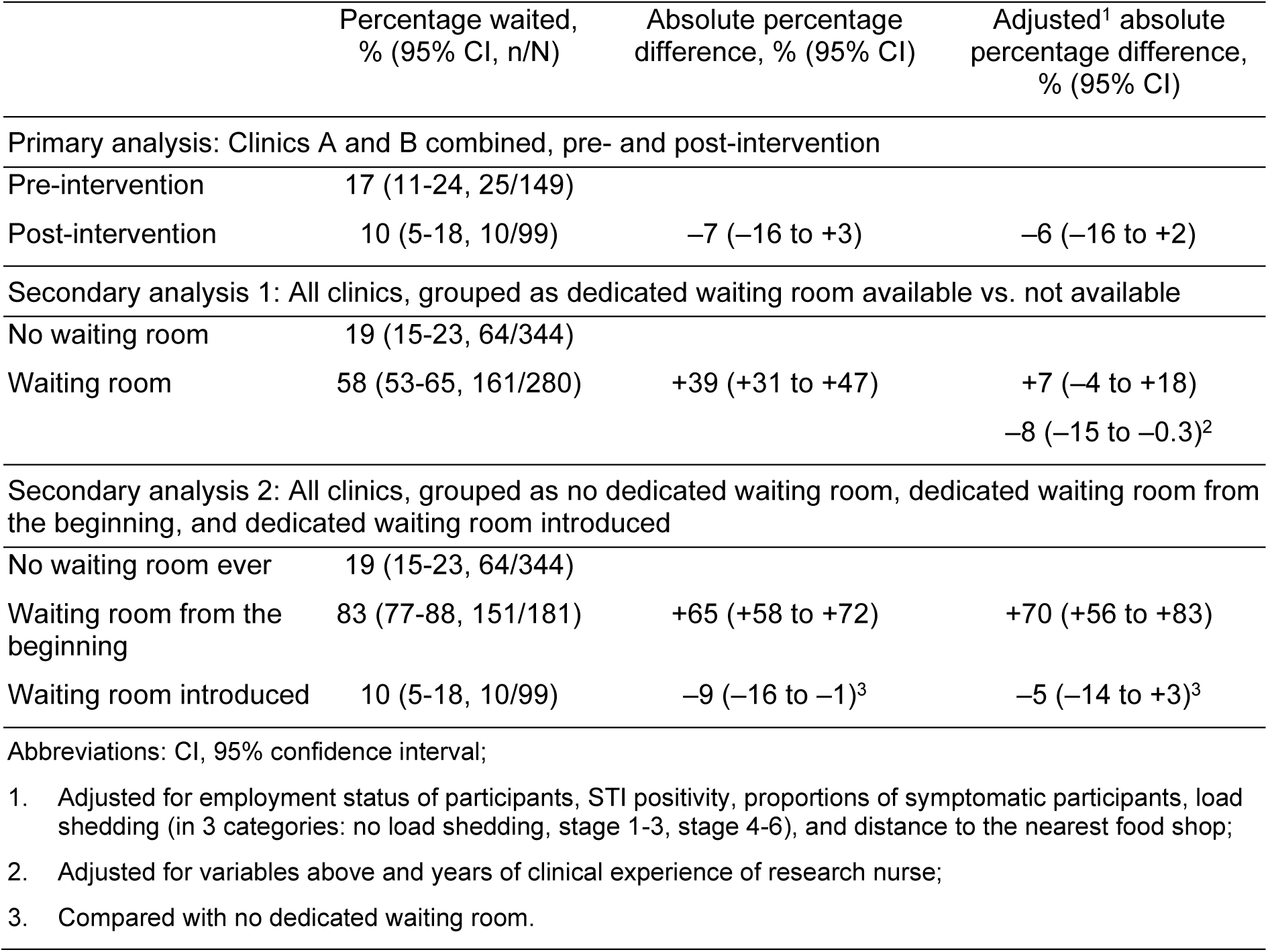
Percentage of women who waited for results, primary and secondary analyses.

For the secondary analysis, with waiting room as a binary variable, 58% (CI 53, 65) of participants waited for their result when a waiting room was available and 19% (CI 15 to 23) when there was no waiting room (absolute difference 39%, CI 31 to 47). After adjustment for employment status, STI test positivity, symptoms, load shedding and distance to the nearest shop, the adjusted percentage difference was +7% (CI –4 to +18) when a waiting room was available (Table 3). When the clinical experience of the nurse was added to the model, the adjusted percentage difference was –8% (CI –15 to –0.3) (Additional File 3).

With waiting room as a 3-level categorical variable, the percentages of participants waiting were: no dedicated waiting room ever 19% (CI 15 to 23), waiting room available from the beginning 83% (CI 77 to 88), and dedicated waiting room introduced 10% (CI 5, to18). Compared with no dedicated waiting room ever, the absolute percentage difference was +65% (CI +58 to +72) when a waiting room was available from the beginning and –9% (CI – 16 to –1) when a waiting room was introduced. In the multivariable model, distance to the nearest shop and clinical experience of the research nurse were highly correlated and could not both be included. After adjustment for employment status of participants, STI positivity, symptoms, load shedding, and distance to the nearest shop, the adjusted percentage differences waiting were similar to the univariable analysis (Table 3).

In the subgroup analysis, according to the presence of an STI, we used data for Clinics A and B. We did not find evidence for an interaction of the STI status and waiting (p-value for interaction unadjusted = 0.79, p-value for interaction adjusted = 0.76, Additional File 4).

Three hundred and ninety-two of 624 (63%) participants provided data about intentions and reasons for not waiting for results (Table 4). Responses did not differ substantially by clinic (Additional File 5) and 136/392 (35%) were collected before the intervention. Of these, 55% (217/392) reported that they did not intend to wait for their results. The main reasons were family (30%, 66/217) or work/school (28%, 60/217) commitments. When we asked these 217 participants what would change their mind about waiting, 72% (157/217) reported that nothing would change their mind. Of 11 participants who did not intend to wait but who received same-day results, 7 said that they were still waiting for other procedures at the clinic, for example for medication from the pharmacy (Table 4). Of 175 participants who intended to wait for their results, 77% (103/175) waited.

**Table 4.** Reasons for not waiting for STI test results, or for changing intention.

| "What is your main reason why you are not intending to wait today?" | Overall (n=217), n (%) |
| --- | --- |
| Have to get back to my kids/family | 66 (30) |
| Have to get to work/school | 60 (28) |
| Hungry | 29 (13) |
| Want to go to the shop | 26 (12) |
| Not feeling well | 11 (5) |
| No time/going somewhere | 8 (4) |
| Load shedding | 5 (2) |
| Referred to hospital | 4 (2) |
| Too hot | 2 (1) |
| Boring | 2 (1) |
| Transport availability | 1 (0.5) |
| No space to wait | 1 (0.5) |
| Didn't come alone to the clinic | 1 (0.5) |
| Tired | 1 (0.5) |
| "What would make you change your mind?" | Overall (n=217), n (%) |
| Nothing | 157 (72) |
| If I did not have other commitments/not be in a hurry | 26 (12) |
| Food | 23 (11) |
| If I had someone looking after the kids | 6 (3) |
| If I can feel better | 3 (1) |
| Comfortable waiting space | 2 (1) |
| "What made you change your mind about waiting for the results?" | Overall (n=11), n (%) |
| Waiting for other clinic procedures (for example medication) | 7 (64) |
| Education about importance of waiting by the nurse | 3 (27) |
| Returned to clinic after being called | 1 (9) |

## Discussion

The absolute difference in Clinics A and B after the introduction of a waiting room was –7% (–16 to +3) in univariable analysis and –6% (–16 to +2) in multivariable analysis. The percentages of pregnant women waiting for STI test results were higher throughout the study period in Clinics C and D, which always had a dedicated waiting room, than in intervention Clinics A and B, and in Clinic E, which never had a dedicated waiting room. Most participants overall (217/392, 55%) said that they were not willing to wait for their results and of those not intending to wait, 72% (157/217) stated that nothing would change their mind.

### Strengths and limitations of the study methods

The main strength of this healthcare improvement study is the real-world setting in multiple primary care clinics, which allowed comparisons both pre- and post-intervention and between clinics in which the availability of a waiting room differed. The study setting and observational design are methodological limitations because the number of clinics was limited and a randomised or crossover design were not possible. Whilst we pre-specified factors that could have affected the tendency to wait for STI test results, the measured variables might not have fully captured the reasons for differences between the study clinics. Measurement error, for example, resulting from applying national-level data to each clinic, could have resulted in a non-differential error, biasing any effect towards the null. In addition, the multivariable analyses aimed to adjust for differences between clinics, but some variables could not be included because the values were identical between clinics or were highly correlated. We collected data about intentions and reasons for waiting for test results after deciding to evaluate the intervention, so these questions were only presented to a subset of participants, and potentially important reasons might have been missed because the responses were pre-defined.

### Interpretation of the study findings

Following installation of a dedicated waiting room, water dispenser and bathroom in two community health facilities, the percentage of pregnant women waiting for the results of on-site GeneXpert tests for STIs did not increase. It was plausible that upgrading the physical infrastructure of research study clinics could have made it more comfortable for women to wait for their results because we had observed much higher percentages of women waiting for results in a healthcare facility with a dedicated waiting room than in clinics without [13]. There are several possible reasons for the primary findings observed in our study. First, the presence or absence of a waiting room in itself might not be a decisive factor influencing decisions about waiting for results. Responses from a subset of participants in this study (Table 4) support this, as very few women said that the absence of a waiting room was the main reason for not waiting and availability of a waiting room would not have changed the decision. This information was only obtained, however, after the decision to install dedicated waiting rooms had been made. Second, an increase in load shedding during the study period might have masked any possible effect of the new waiting rooms. The percentage of women waiting was lower after the intervention than before; power cuts would have decreased the capacity of the GeneXpert machines, made the clinic environment less conducive to waiting and further prolonged clinic visits. Third, there is imprecision in the estimates, which are compatible with both a reduction and a small increase in the percentage of participants waiting.

In other research studies in resource-limited settings, 90% or more of pregnant women have been reported to wait for same-day results and/or treatment following GeneXpert STI testing [9-12]. In our study, similar percentages of women waited for results in some clinics. The large differences between clinics likely reflect a mix of setting-related factors, which we could not entirely disentangle. These clinics differed in other ways from the clinics at which few women waited; locations were in less densely populated areas and closer to the nearest shop, and research nurses were more experienced. In our secondary analysis, the crude percentage difference was 39% (95% CI 31 to 47) higher when a waiting room was available than when it was not available (Table 3). This difference was reduced to +7% (95% CI –4 to +18) after adjusting for factors, including the distance to the nearest shop. This variable may represent other unmeasured community-related characteristics associated with ability to wait for test results. Additional adjustment for the experience of the research nurse reduced this difference further to –8% (95% CI –15 to –0.3) (Additional File 3). The duration of clinical experience of research nurses might therefore also influence outcomes or might represent other unmeasured factors, which could be investigated in future qualitative studies. It is also possible that the physical environment of freight containers and their placement away from the main clinic building (Figure 1) was less attractive than fixed integrated spaces within a healthcare facility.

### Implications for research and policy

This healthcare improvement study shows both the importance and the challenges of evaluating the outcomes of changing physical conditions in the healthcare setting. The goal of improving healthcare infrastructure through initiatives such as the ‘Ideal Clinic’ is clearly important [14] but understanding the impact on process and outcome indicators is also needed. Relevant modifiable factors include clinic workflow, staff training, clinic infrastructure and the turnaround time of test technology. Point-of-care tests that allow treatment decisions to be made at the initial clinical encounter and are rapid, affordable, electricity-independent are particularly important in settings with limited healthcare infrastructure [4-6, 24]. Currently licensed platforms, such as GeneXpert, for on-site *C. trachomatis*, *N. gonorrhoeae* and *T. vaginalis* detection, remain restricted to research studies in most settings. Long waiting times for GeneXpert STI test results have been reported as a major reason for not waiting, in a study amongst youth in Zimbabwe [7] and interruptions to the supply of electricity also reduce efficiency and limit the ability to reduce overall turnaround times. Both qualitative and quantitative studies will be needed to evaluate reasons for differences in, and the implementation of interventions to improve the probability of, waiting for STI results.

## Conclusion

Changing only the physical infrastructure by introducing a dedicated and comfortable waiting room to primary healthcare facilities did not overcome the challenges of providing same-day treatment among pregnant women in our study setting. Future studies should investigate how infrastructure, individual and test-based factors, including other methods of results notification or point-of-care tests with faster turnaround times can improve the uptake of same-day STI testing and treatment.

## Data Availability

All data produced in the present study are available upon reasonable request to the authors

## List of abbreviations

CI: confidence interval
REDCap: Research Electronic Data Capture
STI: sexually transmitted infections

## List of additional files

Additional file 1: Additional file 1.docx, Socio-economic and health characteristics of included and excluded participants.

Additional file 2: Additional file 2.docx, Percentage of women who waited for results by clinic and intervention date (14 December 2023).

Additional file 3: Additional file 3.docx, Secondary analysis 1: All clinics, grouped by waiting room availability.

Additional file 4: Additional file 4.docx, Additional file 4 – Sub-group analysis: Clinics A and B combined, grouped by presence of a sexually transmitted infection.

## Declarations

### Consent for publication

Not applicable

### Availability of data and materials

The datasets analysed for this study are part of ongoing studies, which have not yet been completed. Data used in these analyses are available from the corresponding author on reasonable request.

### Authors’ contributions

RMSG, MMM, NL and RPHP conceptualised the idea and design of this study. RMSG, MMM and RPHP planned and executed data acquisition. RMSG, LB, NL and RPHP performed the statistical analysis. RMSG and NL drafted the manuscript. NL, RPHP, JDK and AMM supervised the study. All authors contributed to interpretation of the data, commented on drafts of the manuscript and approved the final version.

## Acknowledgements

The authors thank all the participants and our field staff.

## Footnotes

1 The alphabetical labelling of the clinics differs from Mdingi et al.’s paper [13].

