## Additional file for "Does a waiting room increase same-day treatment for sexually transmitted infections among pregnant women? A quality improvement study at South African primary healthcare facilities"

### Additional files 1 to 5

### Additional files 1 to 5

#### Additional file 1 – Socio-economic and health characteristics of included and excluded participants

| | Included (N=624) | Excluded (N=876) | P-value |
|---|---|---|---|
| **Age (in years)** | | | |
| Mean (SD) | 27.6 (6.20) | 29.0 (5.96) | <0.001 |
| Median [Min, Max] | 27.0 [18.0, 44.0] | 29.0 [17.0, 44.0] | |
| **Gestational age (in weeks)** | | | |
| Mean (SD) | 14.4 (5.86) | 14.2 (5.82) | 0.366 |
| Median [Min, Max] | 14.1 [3.43, 26.9] | 13.7 [2.86, 26.9] | |
| Missing | 0 (0%) | 4 (0.5%) | |
| **Number of previous pregnancies** | | | |
| Mean (SD) | 1.34 (1.27) | 1.42 (1.29) | 0.266 |
| Median [Min, Max] | 1.00 [0, 10.0] | 1.00 [0, 7.00] | |
| **Education level** | | | |
| Less than Grade. 10 | 48 (7.7%) | 55 (6.3%) | 0.02 |
| Grade 10 or 11 | 209 (33.5%) | 271 (30.9%) | |
| Grade 12 | 310 (49.7%) | 419 (47.8%) | |
| Diploma | 38 (6.1%) | 89 (10.2%) | |
| Degree | 19 (3.0%) | 41 (4.7%) | |
| Missing | 0 (0%) | 1 (0.1%) | |
| **Employment status** | | | |
| Not employed | 383 (61.4%) | 501 (57.2%) | 0.116 |
| Employed | 241 (38.6%) | 375 (42.8%) | |
| **Personal income (ZAR per month)** | | | |
| None | 383 (61.4%) | 505 (57.6%) | 0.001 |
| < 1000 | 31 (5.0%) | 21 (2.4%) | |
| 1001 - 5000 | 157 (25.2%) | 249 (28.4%) | |
| 5001 - 10'000 | 48 (7.7%) | 75 (8.6%) | |
| > 10'000 | 5 (0.8%) | 26 (3.0%) | |
| **Vaginal discharge (observed or reported)** | | | |
| Yes | 82 (13.1%) | 106 (12.1%) | 0.602 |
| No | 542 (86.9%) | 770 (87.9%) | |
| **HIV status** | | | |
| HIV negative | 433 (69.4%) | 615 (70.2%) | 0.833 |
| HIV positive on ART | 158 (25.3%) | 210 (24.0%) | |
| HIV positive, not on ART | 33 (5.3%) | 49 (5.6%) | |
| Missing | 0 (0%) | 2 (0.2%) | |
| **Time normally spent waiting to see a nurse/doctor (in minutes)** | | | |
| Mean (SD) | 62.2 (55.6) | 56.0 (50.3) | 0.123 |
| Median [Min, Max] | 45.0 [0, 360] | 45.0 [0, 420] | |
| **Travelling time to the clinic (in minutes)** | | | |
| Mean (SD) | 25.4 (17.6) | 23.9 (14.0) | 0.797 |
| Median [Min, Max] | 20.0 [0, 120] | 20.0 [1.00, 120] | |
| **Money spent to attend clinic (in ZAR)** | | | |
| Mean (SD) | 37.4 (34.8) | 39.2 (35.8) | 0.651 |
| Median [Min, Max] | 34.0 [0, 300] | 32.0 [0, 300] | |

### Additional files 1 to 5

#### Additional file 2 – Percentage of women who waited for results by clinic and intervention date (14 December 2023)

| Waited for results, % (95% CI) n/N | Clinic A | Clinic B | Clinic C | Clinic D | Clinic E | Overall |
|---|---|---|---|---|---|---|
| | Intervention: waiting room introduced | | No intervention: always had a waiting room | | No intervention: never had a waiting room | |
| Pre-intervention | 26 (16-39)<br>17/65 | 10 (4-18)<br>8/84 | 94 (86-98)<br>77/82 | 68 (43-86)<br>13/19 | 18 (12-26)<br>21/118 | 37 (32-42)<br>136/368 |
| Post-intervention | 18 (9-32)<br>9/50 | 2 (0-12)<br>1/49 | 80 (66-90)<br>41/51 | 69 (49-84)<br>20/29 | 23 (15-35)<br>18/77 | 35 (29-41)<br>89/256 |
| Total | 23 (16-32)<br>26/115 | 7 (3-13)<br>9/133 | 89 (82-93)<br>118/133 | 69 (54-81)<br>33/48 | 20 (15-26)<br>39/195 | 36 (32-40)<br>225/624 |

#### Additional file 3 – Secondary analysis 1: All clinics, grouped by waiting room availability

| | Waiting room vs. no waiting room |
|---|---|
| Absolute percentage difference,<br>% (95% confidence interval) | +39 (+31 to +47) |
| Adjusted absolute percentage difference,<br>% (95% confidence interval) | |
| Adjusted for employment status of participants | +39 (+31 to +47) |
| + STI positivity | +41 (+33 to +49) |
| + proportions of symptomatic participants | +39 (+31 to +47) |
| + load shedding<br>(in 3 categories: no load shedding, stage 1-3, stage 4-6) | +42 (+35 to +50) |
| + the distance in metres to the nearest food shop | +7 (–4 to +18) |
| + work experience of the nurse<br>(in 3 categories: 9 years=low, 15 years=middle, 33 years=long) | –8 (–15 to –0.3) |

### Additional files 1 to 5

#### Additional file 4 – Sub-group analysis: Clinics A and B combined, grouped by presence of an STI

| | Percentage waited, % (95% CI, n/N) | Absolute percentage difference, % (95% CI) | Adjusted[1] absolute percentage difference, % (95% CI) |
|---|---|---|---|
| No STI | 13 (7-21, 13/101) | | |
| Pre-intervention | 18 (7-35, 6/34) | | |
| Post-intervention | 10, (4-20, 7/70) | –9 (–26 to –7) | –11 (–29 to +7) |
| STI present | 15 (10-22, 22/147) | | |
| Pre-intervention | 16 (10-24, 19/118) | | |
| Post-intervention | 10 (3-29, 3/29) | –6 (–20 to –8) | –5 (–23 to +12) |
| p-value for interaction between STI presence and absence | | 0.79 | 0.76 |

Legend: CI, 95% confidence interval; 1, adjusted for employment status of participants, STI positivity, proportions of symptomatic participants, load shedding (in three categories: no load shedding, stage 1-3, stage 4-6), and the distance in metres to the nearest food shop

### Additional files 1 to 5

#### Additional file 5 – Reasons for not waiting for STI test results, or for changing intention, grouped by clinic and pre/post-intervention

| | Clinic A | Clinic B | Clinic C | Clinic D | Clinic E | Overall |
|---|---|---|---|---|---|---|
| Total number of participants | 115 | 133 | 133 | 48 | 195 | 624 |
| Responded to questions about waiting for results, n (%) | 74 (64) | 77 (58) | 76 (58) | 40 (83) | 125 (64) | 392 (63) |
| **"Are you planning to wait for your results today?"** | | | | | | |
| Yes | 48 (65) | 7 (9) | 65 (86) | 35 (88) | 20 (16) | 175 (45) |
| No | 26 (35) | 70 (91) | 11 (15) | 5 (13) | 105 (84) | 217 (55) |
| **"What is your main reason why you are not intending to wait today?"** | **Clinic A (n=26) n (%)** | **Clinic B (n=70) n (%)** | **Clinic C (n=11) n (%)** | **Clinic D (n=5) n (%)** | **Clinic E (n=105) n (%)** | **Overall (n=217) n (%)** |
| Have to get back to my kids/family | 5 (19) | 13 (19) | 3 (27) | 2 (40) | 43 (41) | 66 (30) |
| Have to get to work/school | 8 (31) | 18 (26) | 5 (46) | 1 (20) | 28 (27) | 60 (28) |
| Hungry | 5 (19) | 12 (17) | 0 | 1 (20) | 11 (11) | 29 (13) |
| Want to go to the shop | 0 | 9 (13) | 1 (9) | 0 | 16 (15) | 26 (12) |
| Not feeling well | 2 (8) | 8 (11) | 0 | 0 | 1 (1) | 11 (5) |
| No time/going somewhere | 0 | 5 (7) | 1 (9) | 0 | 2 (2) | 8 (4) |
| Load shedding | 3 (12) | 2 (3) | 0 | 0 | 0 | 5 (2) |
| Referred to hospital | 1 (4) | 1 (1) | 0 | 0 | 2 (2) | 4 (2) |
| Too hot | 0 | 2 (3) | 0 | 0 | 0 | 2 (1) |
| Boring | 0 | 0 | 0 | 0 | 2 (2) | 2 (1) |
| Transport availability | 0 | 0 | 1 (9) | 0 | 0 | 1 (1) |
| No space to wait | 1 (4) | 0 | 0 | 0 | 0 | 1 (1) |
| Didn't come alone to the clinic | 0 | 0 | 0 | 1 (20) | 0 | 1 (1) |
| Tired | 1 (4) | 0 | 0 | 0 | 0 | 1 (1) |
| **"What would make you change your mind?"** | **Clinic A (n=26) n (%)** | **Clinic B (n=70) n (%)** | **Clinic C (n=11) n (%)** | **Clinic D (n=5) n (%)** | **Clinic E (n=105) n (%)** | **Overall (n=217) n (%)** |
| Nothing | 20 (77) | 28 (40) | 11 (100%) | 3 (60) | 95 (91) | 157 (72) |
| If I did not have other commitments/not be in a hurry | 2 (8) | 23 (33) | 0 | 0 | 1 (1) | 26 (12) |
| Food | 3 (12) | 10 (14) | 0 | 1 (20) | 9 (9) | 23 (11) |
| If I had someone looking after the kids | 0 | 5 (7) | 0 | 1 (20) | 0 | 6 (3) |
| If I can feel better | 0 | 3 (4) | 0 | 0 | 0 | 3 (1) |

### Additional files 1 to 5

| | | | | | | |
|---|---|---|---|---|---|---|
| Comfortable waiting space | 1 (4) | 1 (1) | 0 | 0 | 0 | 2 (1) |

| **"What made you change your mind about waiting for the results?"** | **Clinic A (n=1), n (%)** | **Clinic B (n=0), n (%)** | **Clinic C (n=1), n (%)** | **Clinic D (n=2), n (%)** | **Clinic E (n=7, n (%)** | **Overall (n=11) n (%)** |
|---|---|---|---|---|---|---|
| Waiting for other clinic procedures (for example medication) | 1 (100) | 0 | 0 | 0 | 6 (86) | 7 (64) |
| Education about importance of waiting by the nurse | 0 | 0 | 1 (100) | 2 (100) | 0 | 3 (27) |
| Returned to clinic after being called | 0 | 0 | 0 | 0 | 1 (14) | 1 (9) |
